# The benefit of hypothermic targeted temperature management decreases when post-arrest care improves

**DOI:** 10.1101/2023.05.22.23290369

**Authors:** Wei-Ting Chen, Yu-Tzu Tien, Chien-Hua Huang, Chih-Hung Wang, Wei-Tien Chang, Hooi-Nee Ong, Wen-Jone Chen, Min-Shan Tsai

## Abstract

**Background:** Several clinical trials and cohort studies in the past two decades have demonstrated inconsistent results regarding survival and neurologic recovery benefits for patients undergoing targeted temperature management (TTM) at 33℃ as compared to those undergoing TTM at 36℃ or normothermia. Whether the improved quality of post-arrest care over time contributes to diminishing benefit of TTM at 33℃ remains un-investigated.

**Methods:** There were 1,809 adult non-traumatic cardiac arrest survivors during 2011-2020. After excluding patients with pre-arrest poor conscious level defined as Glasgow-Pittsburgh Cerebral Performance Category (CPC) >2 (n=258), patients who regained clear consciousness after return of spontaneous circulation (ROSC) (n=300), patients with active bleeding or intracranial hemorrhage (n=48) and patients who underwent TTM of 36℃ due to unstable hemodynamic (n=11), a total of 1,192 eligible candidates for TTM of 33℃ were finally enrolled and classified into Period 1 (during 2011-2015, n = 449) and Period 2 (during 2016-2020, n = 693).

**Results:** Patients in Period 2 received more diagnostics procedures and specific therapies than those in Period 1. The proportion of patients with good neurological recovery at hospital discharge in Period 2 is significantly higher than that in Period 1 (21.4% vs.14.0%, adjusted odds ratio [aOR] 1.60, 95% confidence interval [CI] 1.14-2.26). TTM was beneficial for the outcomes in both Period 1 and 2, with patients in Period 1 having higher chances of survival and good neurological outcome than those in Period 2 (survival: aOR 5.66, 95% CI 3.49-9.18 vs. aOR 2.91, 95% CI 1.98-4.28; good neurological recovery: aOR 3.92, 95% CI 2.12-7.25 vs. aOR 2.19, 95% CI 1.43-3.34). Among patients with low-risk and medium-risk severity of illness, TTM benefited survival and neurological outcomes, regardless of time period. But the chance of beneficial outcomes decreased consistently from Period 1 to Period 2.

**Conclusion:** Among cardiac arrest survivors, improvement in the quality of post-cardiac arrest care over time is associated with better neurological recovery. TTM remains beneficial for survival and neurological outcomes following cardiac arrest, regardless of the time period. However, the benefit of TTM may diminish when post-cardiac arrest care improves.

## Background

In recent decade, the dispute whether preventing fever following return of spontaneous circulation (ROSC) rather than reducing body temperature to 33℃ was enough to alleviate post-arrest brain injury has been raised, since the TTM and TTM2 trials demonstrated no difference in survival and neurological outcomes between patients underwent TTM of 33℃ vs. 36℃,^1^ or vs. <37.8℃,^2^. However, several retrospective cohort studies have shown less protocol compliance^3^ and more fever during the early post-arrest period,^3,4^ and thus worse survival and neurological recovery in patients underwent targeted temperature management (TTM) of 36℃.^4–6^ Moreover, the HYPERION trial demonstrated better neurological recovery in TTM of 33℃ than normothermia among patients with non-shockable rhythm.^7^ The severity of post-cardiac arrest syndrome was proposed to account for the discrepancies between these studies. Better outcomes were observed when temperature was targeted at 33℃ in the patients with moderate-severity.^8,9^

In addition to illness severity, the benefit of protocolized post-arrest care in the trials may also attribute to the insignificant difference between hypothermia and normothermia. The importance of post-cardiac arrest care began to be emphasized since 2010, and a multidisciplinary, systemic approach of care was proposed, including optimization of ventilation and hemodynamics, neurologic management, glycemic control, TTM and immediate coronary revascularization.^10,11^ Our previous study demonstrated that these practices help improve the quality of care in patients without TTM.^12^ However, whether benefit of therapeutic hypothermia over no temperature management decreases along with the improved post-arrest care remains uninvestigated. Before the launch of the 2015 American Heart Association (AHA) guidelines, the National Taiwan University Hospital (NTUH) only used the TTM protocol with a target temperature of 33℃. Since 2016 January, the NTUH protocol has been revised to subject stable patients to TTM at 33℃ while only a few patients with unstable hemodynamics or arrhythmia are treated at 36℃. The aim of this study was to examine the benefit of hypothermic TTM at 33℃ on survival and favorable neurologic recovery during the time period from 2010 to 2020, in the context of the improvements of post-arrest care.

## Methods

### Demographics/Epidemiology

This retrospective cohort study was conducted at National Taiwan University Hospital, a 2500-bed tertiary medical center located in Taipei City with 110,000 annual emergency department visits^13^. The Institutional Review Board of NTUH approved the study(202111079RINA) and waived participant consent.

#### Participants Selection

The study enrolled 1,809 adult, non-traumatic cardiac arrest victims with successful resuscitation between January 2011 and December 2020 in the ED of NTUH. Patients were excluded if they had a pre-arrest poor consciousness level, defined as Glasgow-Pittsburgh Cerebral Performance Category (CPC)>2 (n=258), had regained clear consciousness within 3 hours after ROSC (n=300), had massive bleeding or intracranial hemorrhage at ROSC (n=48), and who underwent TTM with a target temperature of 36℃ due to unstable hemodynamics (n=11). A total of 1,192 eligible candidates for TTM with target temperature at 33℃ were finally enrolled in the current study. When further classified by the AHA guidelines revised in Dec 2015, 449 patients with an index cardiac arrest between 2011-2015 were further classified as “Period 1”, and 693 patients with an index cardiac arrest between 2016-2020 were classified as “Period 2” (Figure 1).

**Figure 1.**
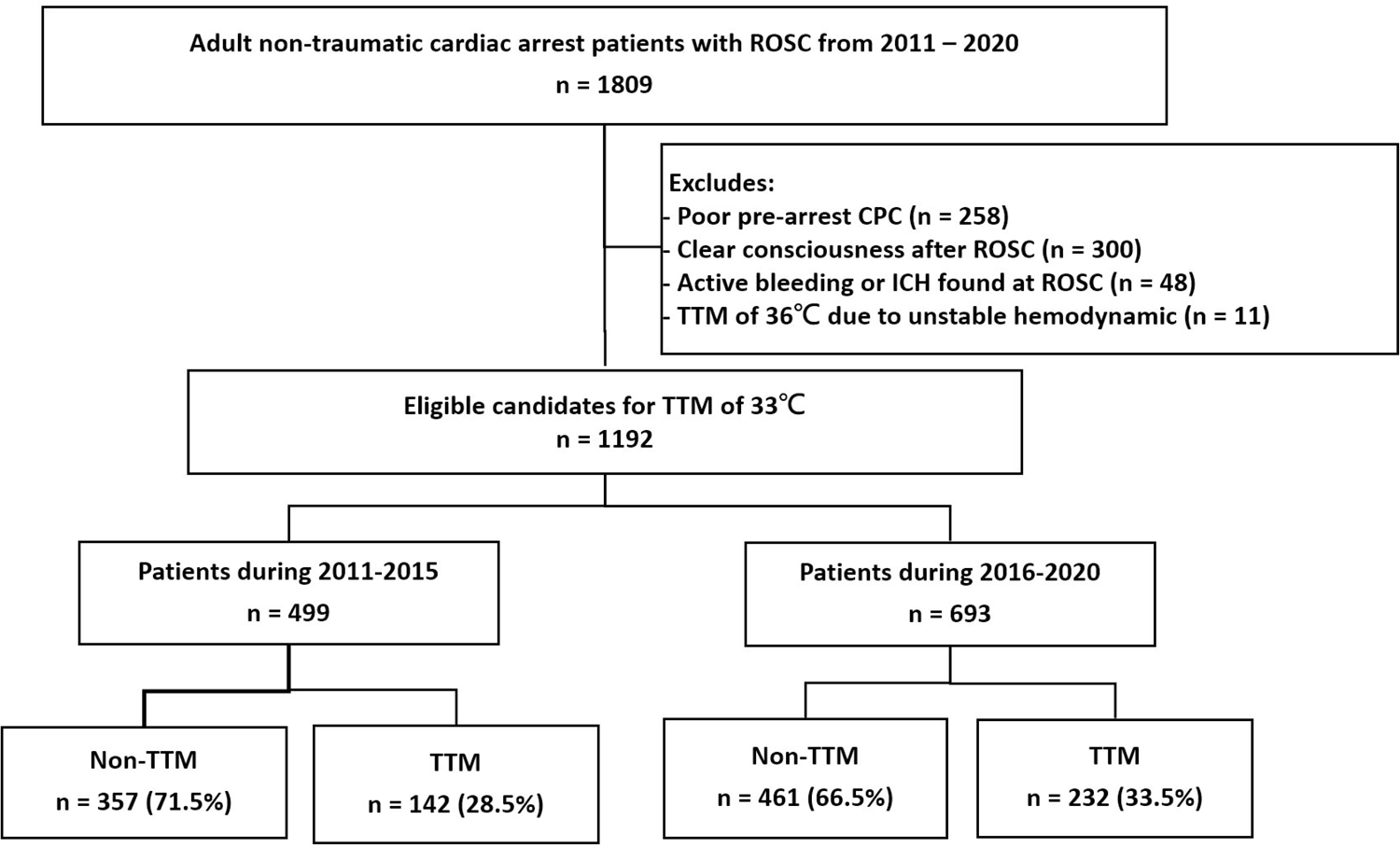
The flowchart of enrolled patients. CPC, Glasgow-Pittsburgh Cerebral Performance Category; ICH, intracranial hemorrhage; ROSC, return of spontaneous circulation; TTM, targeted temperature management.

#### TTM Protocol of NTUH

NTUH is the first hospital in Taiwan to introduce TTM for cardiac arrest survivors, and has established a protocol for TTM since 2003. After the application of cooling devices was approved by the Taiwan Ministry of Health and Welfare in 2005, the first patient underwent TTM in 2006. The protocols for post-cardiac arrest care, including TTM, were updated sequentially in alignment with the 2010, 2015 and 2020 AHA guidelines.^10,14,15^ Before the launch of 2020 AHA guideline, the cardiac arrest survivors underwent TTM at 33°C except for those with unstable hemodynamic even with more than 2 inotropes support in the early post-arrest period and those with massive bleeding or intracranial hemorrhage after ROSC. Cold saline and cooling devices with auto-feedback were used to reduce patients’ body temperatures to the target temperature of 33°C within 4–6 hours after ROSC, maintain target temperature for 24 hours, and rewarm patients at the rate of 0.25°C per hour until 36°C was achieved. Temperature management was continued for another 24 hours after rewarming to avoid fever ^13^.

#### Data Collection and Variables Definition

The primary data set was obtained from the hospital medical record data repository and included demographic information, past medical history, prehospital arrest events, cardiac arrest details, post-arrest management, laboratory examinations, and disposition. This study followed Strengthening the Reporting of Observational Studies in Epidemiology (STROBE) reporting guidelines.^16^

Patients with cardiac arrest were considered as either those with out-of-hospital cardiac arrest (OHCA), with in-hospital cardiac arrest (IHCA) after triage, or those who achieved ROSC before being transferred from another hospital. Cardiogenic arrest was recorded when ischemic heart disease, structural heart disease, heart failure, or arrhythmia without an electrolyte imbalance was considered the cause of arrest. The causes of cardiac arrest were determined by the responsible primary care physicians, who were blinded to the current study. Repeated CPR was defined as another episode of arrest within one hour after initial ROSC.

Overall cardiac arrest severity was assigned using the simplified Cardiac Arrest Hospital Prognosis (sCAHP) score as our previous publication, which excluded the no-flow interval data with similar predictive accuracy as the original score in an East Asian population.^17^ The illness severity (≤150 as low-risk, 151-199 as medium-risk, and ≥200 as high-risk) was determined by using the method with a scheme that used significant F-values for cut-off points.^18^ The performance of illness severity categorized by the sCAHP score (our data) and the CAHP score (Sudden Death Expertise Center registry, Paris, France)^19^ is shown in Supplementary Table 1.

During the early post-arrest period, electrocardiogram (ECG), echocardiography and cerebral computed tomography after ROSC were recorded. Emergent coronary angiography (CAG) was recorded when indicated patients underwent CAG within 24 hours after ROSC. Extracorporeal membrane oxygenation (ECMO) was recorded when applied as indicated during index resuscitation. Fever (≥38.3°C), infection workup, electroencephalogram (EEG) and anti-epileptic drugs (AED) use during first 3 days after ROSC were documented.

#### Outcome Measures

The primary outcome was favorable neurological recovery at hospital discharge, defined as a CPC of 1 (good recovery) or 2 (moderate disability). The secondary outcomes include survival at hospital discharge, as well as at 3-month and 1-year follow-ups. CPC at hospital discharge was evaluated by primary care physicians who were blinded to the current study.

### Patient and public involvement

The patients and public were not involved in the creation of the study design, recruitment or statistical analysis. Patients were not consulted to develop patient relevant outcomes or interpret the results. Patients were not invited to contribute to the writing or editing of this manuscript.

#### Statistical Analysis

Descriptive statistics were presented as frequencies for categorical variables and the median with interquartile ranges for continuous variables. For 2 group comparison, Fisher’s exact or Pearson’s chi-squared test for categorical variables, and the Mann-Whitney U test for continuous variables were used. Multiple imputation (Markov Chain Monte Carlo) was used for missing data of laboratory examinations. Multivariable logistic regression models were used to examine the association between outcomes and time periods, and the benefit of hypothermic TTM over non-TTM on survival and good neurological recovery. All statistical analyses were performed using SPSS Statistics for Windows, version 16.0 (SPSS Inc., Chicago, IL, USA). Statistical significance was assessed using two-sided 95% confidence intervals (CI) and a p<0.05.

## Results

### Period 1 vs. Period 2

A comparison of the patient characteristics, cardiac arrest events, and laboratory examinations between the two time periods is presented in Table 1. As compared with Period 1, the proportion of coronary artery disease, arrhythmia, OHCA, witnessed collapse, initial shockable rhythm and bystander CPR were higher during Period 2. But the adrenaline dosage, systolic blood pressure at ROSC, PH and HCO_3_ of blood gas were lower in patients during Period 2 than Period 1. Patients in Period 2 had higher sCAHP severity and APACHE II scores than those in Period 1. More patients in Period 2 received echocardiography, brain CT, emergent CAG, TTM and inotrope support following ROSC as well as EEG and AEDs within 3 days after cardiac arrest. Patients in Period 1 had longer duration from cooling to target temperature and greater highest body temperature when compared with those in Period 2. Increased active withdrawal was also noted in Period 2 (Table 2).

**Table 1.** Comparison of patient characteristics between time periods.

|  | Overall | Period 1<br>2011-2015 | Period 2<br>2016-2020 | *p-value* |
| --- | --- | --- | --- | --- |
|  | **n = 1192** | **n = 499** | **n = 693** |  |
| Male Sex (%) | 802 (67.3) | 325 (65.1) | 477 (68.8) | 0.189 |
| Age, median(IQR), yr | 68 (56-79) | 67 (55-79) | 68 (57-79) | 0.290 |
| Age ≥65 yrs (%) | 693 (58.1) | 276 (55.3) | 417 (60.2) | 0.096 |
| **Underlying Characteristics** |  |  |  |  |
| Hypertension (%) | 622 (52.2) | 244 (48.9) | 378 (54.5) | 0.060 |
| Diabetes mellitus (%) | 365 (30.6) | 155 (31.1) | 210 (30.3) | 0.799 |
| Hyperlipidemia (%) | 112 (9.4) | 37 (7.4) | 75 (10.8) | 0.056 |
| CAD (%) | 346 (29.0) | 123 (24.6) | 223 (32.2) | 0.005 |
| Heart failure (%) | 118 (9.9) | 44 (8.8) | 74 (10.7) | 0.326 |
| VHD (%) | 28 (2.3) | 8 (1.6) | 20 (2.9) | 0.177 |
| Arrhythmia (%) | 141 (11.8) | 48 (9.6) | 93 (13.4) | 0.046 |
| COPD/Asthma (%) | 100 (8.4) | 43 (8.6) | 57 (8.2) | 0.833 |
| Renal disease (%) | 116 (9.7) | 53 (10.6) | 63 (9.1) | 0.428 |
| ESRD (%) | 107 (9.0) | 40 (8.0) | 67 (9.7) | 0.356 |
| CVA (%) | 104 (8.7) | 37 (7.4) | 67 (9.7) | 0.178 |
| Dementia (%) | 50 (4.2) | 25 (5.0) | 25 (3.6) | 0.244 |
| Bedridden (%) | 49 (4.1) | 14 (2.8) | 35 (5.1) | 0.056 |
| Malignancy (%) | 289 (24.2) | 122 (24.4) | 167 (24.1) | 0.891 |
| **Cardiac Arrest Events** |  |  |  |  |
| Source (%) |  |  |  |  |
| OHCA | 736 (61.7) | 285 (57.1) | 451 (65.1) | 0.017 |
| IHCA | 386 (32.4) | 179 (35.9) | 207 (29.9) | — |
| Transfer | 70 (5.9) | 35 (7.0) | 35 (5.1) | — |
| Witnessed collapse (%) | 969 (81.3) | 391 (78.4) | 578 (83.4) | 0.029 |
| Initial shockable rhythm (%) | 289 (24.2) | 99 (19.8) | 190 (27.4) | 0.003 |
| Bystander CPR (%) | 667 (56.0) | 246 (49.3) | 421 (60.8) | <0.001 |
| Prehospital ROSC (%) | 106 (9.6) | 39 (8.8) | 67 (10.0) | 0.533 |
| Adrenaline ≥3 mg (%) | 607 (50.9) | 275 (55.1) | 332 (47.9) | 0.016 |
| CPR duration, median(IQR), min | 19 (9-33) | 18 (9-30) | 20 (9-34) | 0.129 |
| CPR ≤10 min (%) | 338 (29.0) | 142 (29.6) | 196 (28.6) | 0.331 |
| CPR 10-20 min (%) | 251 (21.5) | 112 (23.3) | 139 (20.3) | — |
| CPR ≥20 min (%) | 576 (49.4) | 226 (47.1) | 350 (51.1) | — |
| Cardiogenic arrest (%) | 591 (49.6) | 239 (47.9) | 352 (50.8) | 0.348 |
| Repeated CPR (%) | 409 (34.3) | 161 (32.3) | 248 (35.8) | 0.216 |
| ROSC HR, median(IQR), bpm | 105 (79-128) | 107 (79-129) | 104 (79-129) | 0.573 |
| ROSC SBP, median(IQR), mmHg | 117 (86-152) | 121 (90-159) | 116 (85-151) | 0.039 |
| sCAHP severity (%) |  |  |  |  |
| Low-Risk | 248 (20.8) | 105 (21.0) | 143 (20.6) | 0.002 |
| Medium-Risk | 547 (45.9) | 255 (51.1) | 292 (42.1) | — |
| High-Risk | 397 (33.3) | 139 (27.9) | 258 (37.2) | — |
| APACHE II score, median(IQR) | 32 (26-37) | 28 (24-33) | 34 (30-39) | <0.001 |
| **Laboratory Exams during CPR** |  |  |  |  |
| WBC, median(IQR), K/uL | 12.02<br>(8.94-16.36) | 12.28<br>(9.06-16.69) | 11.84<br>(8.85-16.18) | 0.353 |
| Hemoglobin, median(IQR), g/dL | 12.3 (10.0-14.6) | 12.1 (10.0-14.4) | 12.5 (10.0-14.7) | 0.284 |
| Platelet, median(IQR), K/uL | 190 (139-252) | 187 (134-244) | 194 (142-257) | 0.157 |
| Lactic acid, median(IQR), mmol/L | 9.7 (6.5-13.0) | 9.3 (6.1-12.7) | 9.9 (6.7-13.2) | 0.122 |
| pH, median(IQR), | 7.11 (6.99-7.23) | 7.14 (7.05-7.24) | 7.08 (6.95-7.21) | <0.001 |
| HCO<sub>3</sub>, median(IQR), mmol/L | 19.0 (15.0-23.6) | 19.7 (15.7-24.9) | 18.5 (14.6-22.9) | <0.001 |

Data presented as no. (%) or as median (IQR).
CAD, coronary artery disease; CPR, cardiopulmonary resuscitation; COPD, chronic obstructive pulmonary disease; CVA, cerebrovascular accident; ESRD, end-stage renal disease; HR, heart rate; IHCA, in-hospital cardiac arrest; OHCA, out-of-hospital cardiac arrest; ROSC, return of spontaneous circulation; SBP, systolic blood pressure; sCAHP, simplified Cardiac Arrest Hospital Prognosis; VHD, valvular heart disease.

**Table 2.** Comparison of post-arrest care between time periods.

|  | Overall | Period 1<br>2011-2015 | Period 2<br>2016-2020 | *p-value* |
| --- | --- | --- | --- | --- |
|  | n = 1192 | n = 499 (41.9%) | n = 693 (58.1%) |  |
| ECG after ROSC (%) | 1111 (93.2) | 458 (91.8) | 653 (94.2) | 0.103 |
| Echocardiography after ROSC (%) | 583 (48.9) | 227 (45.5) | 356 (51.4) | 0.046 |
| Brain CT after ROSC (%) | 782 (65.6) | 279 (55.9) | 503 (72.6) | <0.001 |
| Emergent CAG (%) | 250 (21.0) | 85 (17.0) | 165 (23.5) | 0.005 |
| TTM of 33°C (%) | 374 (31.4) | 142 (28.5) | 232 (33.5) | 0.067 |
| ROSC to start cooling, median(IQR), hr | 4 (2-6) | 3 (2-6) | 4 (2-6) | 0.514 |
| Cooling to target temperature, median(IQR), hr | 4 (2-7) | 5 (3-9) | 3 (2-6) | <0.001 |
| Rewarming duration, median(IQR), hr | 14 (12-19) | 15 (11-21) | 14 (12-18) | 0.152 |
| ECMO (%) | 226 (19.0) | 93 (18.6) | 133 (19.2) | 0.823 |
| ROSC 3hr |  |  |  |  |
| MAP≥65 mmHg (%) | 749 (83.8) | 290 (82.4) | 459 (84.7) | 0.403 |
| inotrope use (%) | 751 (63.0) | 293 (58.7) | 458 (66.1) | 0.011 |
| PO2 75-100 mmHg (%) | 138 (11.6) | 59 (11.8) | 79 (11.4) | 0.855 |
| PCO2 35-45 mmHg (%) | 246 (25.9) | 91 (23.5) | 155 (27.5) | 0.176 |
| Lactic acid change from ROSC,<br>median(IQR), mmol/L | -0.90<br>(-6.08-5.20) | -0.16<br>(-5.07-5.46) | -1.64<br>(-6.33-4.45) | 0.067 |
| Decreased Lactic acid from ROSC (%) | 264 (55.7) | 119 (51.1) | 145 (60.2) | 0.052 |
| Within 3 days after ROSC |  |  |  |  |
| Fever(≥38.3°C) (%) | 208 (17.4) | 97 (19.4) | 111 (16.0) | 0.142 |
| Highest body temperature , median(IQR), °C | 37.1<br>(36.0-38.0) | 37.3<br>(36.0-38.1) | 37.0<br>(36.0-37.9) | 0.030 |
| Infection workup (%) | 742 (63.0) | 297 (61.2) | 445 (64.1) | 0.327 |
| EEG (%) | 223 (18.7) | 71 (14.2) | 152 (21.9) | 0.001 |
| AED use (%) | 227 (19.0) | 68 (13.6) | 159 (22.9) | <0.001 |

Data presented as no. (%) or as median (IQR).
AED, anti-epileptic drugs; CAG, coronary angiography; CT, computed tomography; ECG, electrocardiogram; ECMO, extracorporeal myocardial oxygenation; EEG, electroencephalogram; MAP, mean arterial pressure; ROSC, return of spontaneous circulation; TTM, targeted temperature management.

A representation of the proportion of patients that did not survive to hospital discharge, with poor CPC, and with good CPC between the two time periods can be seen in Figure 2. Mortality rates are comparable with 71.1% in Period 1 and 72.2% in Period 2. The chance of patients with good neurological recovery at ICU discharge (adjusted OR [aOR] 1.79, 95% CI=1.27-2.51) and hospital discharge (aOR 1.60, 95% CI=1.14-2.26) in Period 2 was significantly higher than in Period 1. There were no difference of 3-month and 1-year survival between Period 1 and Period 2 (Table 3).

**Figure 2.**
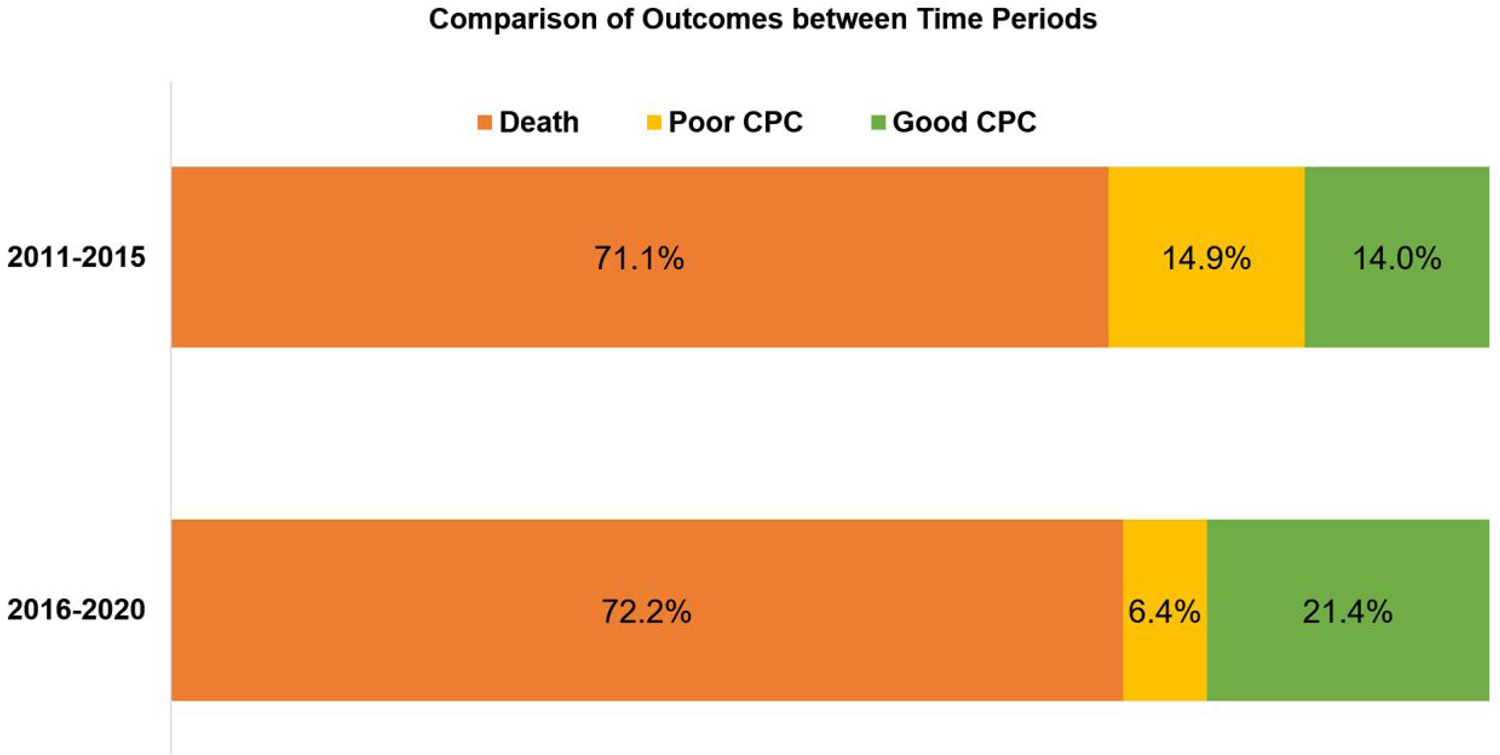
Comparison of Outcomes between Time Periods. CPC, Glasgow-Pittsburgh Cerebral Performance Category

**Table 3.**
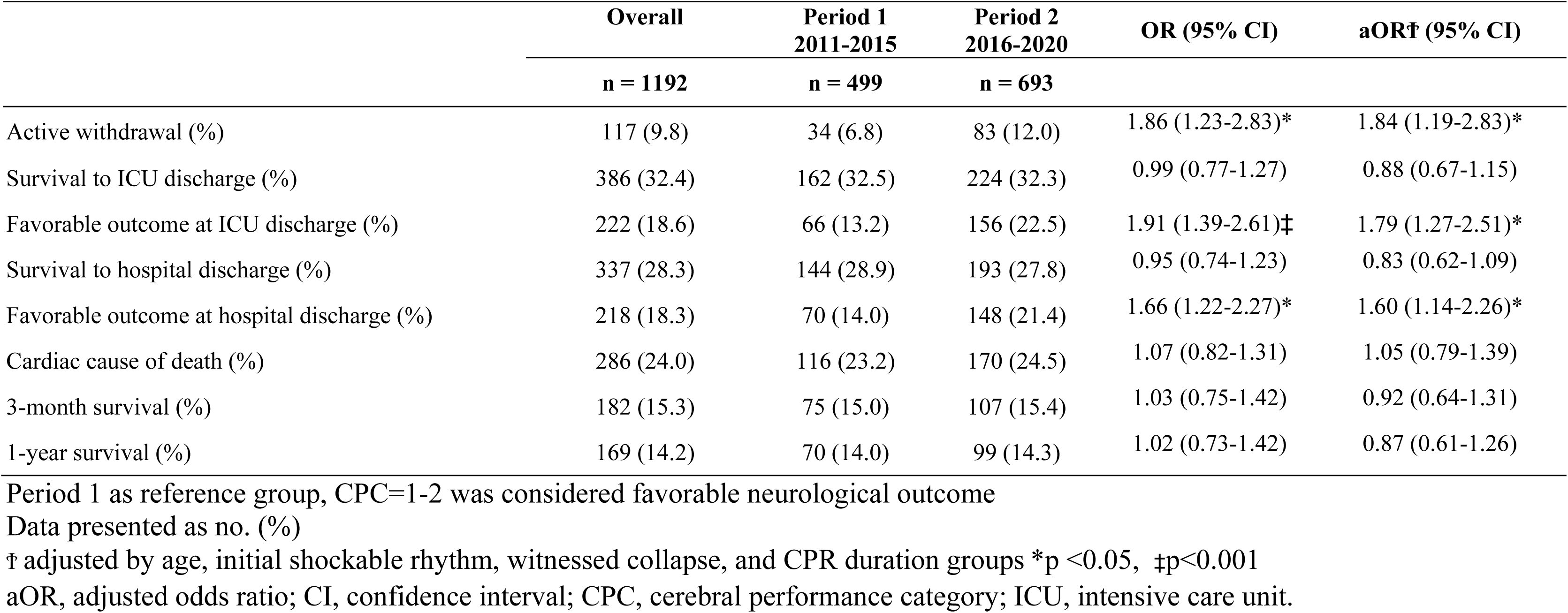
Comparison of outcomes between time periods.

### Non-TTM vs. hypothermic TTM

There were 374 patients receiving hypothermic TTM following ROSC classified as the TTM group and the rest 818 patients classified as the non-TTM group. A comparison of underlying characteristics, cardiac arrest events, and laboratory results between patients with and without TTM treatment are presented in Supplementary Table 2. The patients in the TTM group were younger than those in the non-TTM group and suffered from less pre-arrest co-morbidity, such as bed-ridden and malignancy. Compared with the non-TTM group, the TTM group had a higher proportion of patients with OHCA, initial shockable rhythm, prehospital CPR, prehospital ROSC and presumed cardiogenic arrest, but less percentage of repeated CPR and high-risk severity. Besides, the TTM group required a lower adrenaline dosage, and had lower APACHE II scores but higher heart rates, systolic blood pressures, hemoglobin and platelet concentrations than the non-TTM group. Supplementary Table 3 demonstrates better post-arrest care in the TTM group than the non-TTM group including more cardiovascular and cerebral evaluations, better adherence to ventilation and hemodynamic recommendations, more infection workups, and early seizure detection and treatment. A comparison between the TTM and non-TTM groups in each time period is shown in Supplementary Tables 4 and 5.

Although significant interaction between time period and TTM was noted, both Period 2 and TTM remained associated with improved neurological recovery (Supplementary Table 6). Compared with the non-TTM group, patients who underwent TTM had better survival (aOR 3.67, 95% CI=2.73-4.93) and neurological outcome (aOR 2.71, 95% CI=1.92-3.82). TTM remained beneficial in both Period 1 and Period 2, and the benefit of TTM over non-TTM in survival and neurological recovery was higher in Period 1 as compared to Period 2 (survival: aOR 5.66, 95% CI=3.49-9.18 vs. aOR 2.91, 95% CI=1.98-4.28; good neurological recovery: aOR 3.92, 95% CI=2.12-7.25 vs. aOR 2.19, 95% CI=1.43-3.34). The enrolled patients were further stratified by sCAHP severity with 248 patients of low-risk severity, 547 patients of medium-risk severity and 397 patients of high-risk severity, respectively. Among patients with low- and medium-risk severity, TTM benefited survival and neurological outcomes, regardless of time period. But the chance for good outcomes consistently decreased from Period 1 to Period 2 in both low- and medium-risk subgroups. There was no significant benefit of TTM in patients with high-risk severity (Figure 3).

**Figure 3.**
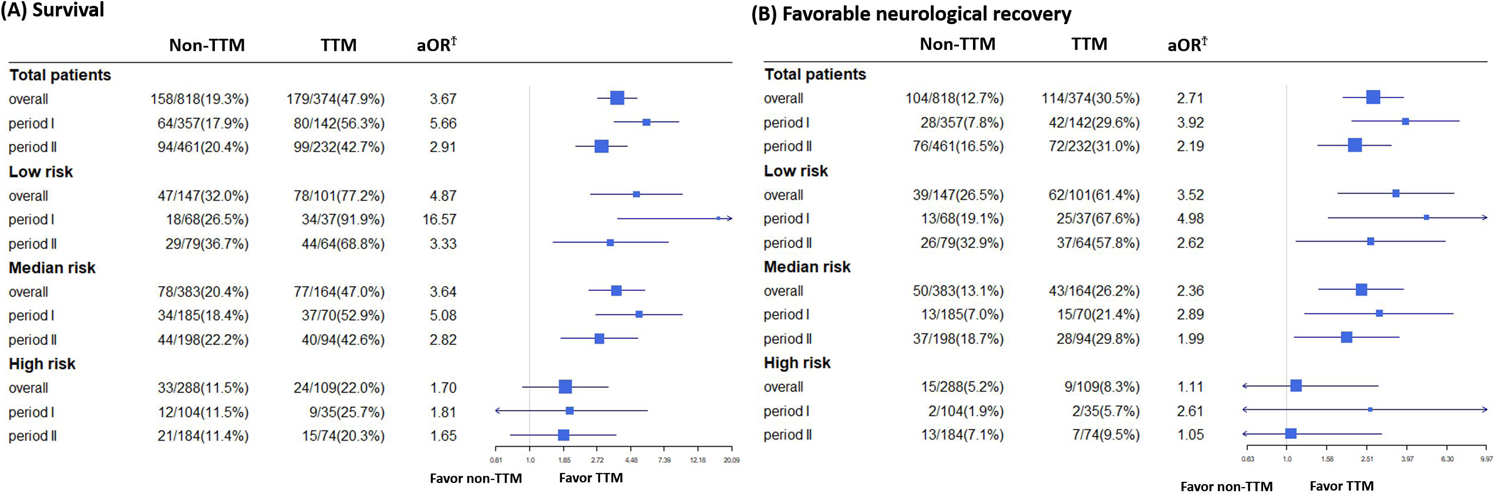
The benefit of TTM over non-TTM in survival and neurological outcomes. Non-TTM as reference group, CPC=1-2 was considered as favorable neurological recovery Ϯ adjusted by age, initial shockable rhythm, witnessed collapse, and CPR duration groups aOR, adjusted odds ratio; CI, confidence interval; CPC, cerebral performance category; TTM, targeted temperature management

## Discussion

In this study, Period 2 had better post-arrest care than Period 1, and the neurological recovery improved significantly in Period 2. TTM were beneficial for survival and neurological outcomes among cardiac arrest patients in both time periods, especially in those with low- and medium-risk severity; however, its benefit diminished over time.

To alleviate post-cardiac arrest syndrome, bundle care focused on identifying the cause of arrest and preventing recurrence, providing ventilation to minimize lung injury, optimizing cardiovascular function, early detecting and treating post-arrest seizure, reducing risk of multiorgan injury, and controlling body temperature has been suggested by AHA guidelines since 2010.^10^ In the current study, patients in Period 2 had increased diagnostics procedures performed and specific therapies given for timely and appropriate management than those in Period 1. Although both short-term and long-term survival are comparable between Period 1 and Period 2, the benefit of improved post-cardiac arrest care might be reflected in more favorable neurological recovery in Period 2. The increased active withdrawal for patients with potentially poor neurological outcome and “do-not-attempt resuscitation” after ROSC in Period 2 may account for the insignificant difference in survival at hospital discharge between these 2 periods. Besides, the finding that approximately half of the patients discharged from the hospital died within 3 months emphasizes the importance of rehabilitation and health care after hospital discharge. Further studies are warrant to investigate the causes of death after hospital discharge.

In our current study, TTM at 33℃ remains beneficial in both time periods. The benefits of hypothermic TTM over non-TTM decreased in Period 2 in all enrolled patients as well as subgroups stratified by illness severity. The improved post-arrest care may account for this finding since the interventions were in place for all successfully resuscitated victims, regardless of TTM treatments. This finding also raises another important question—*Does the benefit of TTM on outcomes truly result from TTM or the protocolized care established along with TTM?* Laboratory animal studies displayed that therapeutic hypothermia at 32-33 °C ameliorated myocardial injury and brain damage after ischaemia/reperfusion injury, and thus, improved survival and neurological recovery.^20,21^ However, clinical trials showed inconsistent results concerning the benefit of TTM at 33 °C as compared with normothermia or no temperature control.^1,2,7,22,23^

Bernard et al., HACA and HYPERION trials demonstrated improved survival and neurological outcomes in patients underwent therapeutic hypothermia at 33 °C.^7,22,23^ However, the TTM trial demonstrated no difference in survival and neurological recovery between patients receiving TTM at 33 °C and 36 °C, emphasizing the importance of temperature control rather than the low target temperature.^1^ In 2021, the TTM2 trial once again demonstrated no outcome benefit in patients receiving TTM at 33 °C compared with those <37.8 °C, implying that aggressively preventing fever may be the main determinant factor.^2^ The evolution of post-cardiac arrest care within the two decades may account for the inconsistent results of these trials. Similar situation can be observed in the care of sepsis. The ARISE trial failed to show the benefit of early goal-directed therapy(EGDT) in reducing mortality, and the improvement of medical care quality along with the protocol establishment was proposed as a reason for comparable outcomes in the EGDT and control groups.^24^ The protocolized approaches usually facilitate medical teams to provide more consistent care and overcome barriers.^25,26^ Our results supported the hypothesis that the improved quality of post-arrest care would diminish the benefit of TTM on outcomes. Several retrospective cohort studies also demonstrated worse outcomes in patients with less adherence to protocols of temperature control. ^3–5^ Similar diminished difference between intervention and standard groups can also be observed in the Box trial: no outcome difference between different blood-pressure targets and different oxygen targets in cardiac arrest survivors. ^27,28^

### Limitation

This study had several limitations. First, due to the its retrospective nature, selection bias was unavoidable, and unidentified confounding factors or improvement of post-arrest care might be present. Second, the post-cardiac arrest care in the TTM group were better than that in the non-TTM group. The frequency of CT, emergent CAG and EEG of the TTM group were comparable with those in published studies.^2,5,9^ However, there is a lack of studies reporting the quality of post-arrest care in patients without TTM and we cannot compare our results with others. Nevertheless, our results indicated that much room for us to improve the care quality in patients without TTM. And the care discrepancy between the TTM and non-TTM groups might be the primary source of the TTM benefit as we hypothesized. Third, the reasons for patients not undergoing TTM were not recorded. The socioeconomic status of these patients, which might affect willingness to undergo TTM, was also not considered. However, standard ICU and post-arrest care are covered by National Health Insurance in Taiwan, and TTM was incorporated into the coverage in October 2015. Additionally, the neurological function was not recorded during the follow-up after hospital discharge. Finally, our study investigated a homogenous Asian population, and great caution should be exercised when attempting to extrapolate our results to other races.

## Conclusion

Among cardiac arrest survivors, the improved quality of post-cardiac arrest care over time is associated with better neurological recovery. Regardless of the time periods, hypothermic TTM remained beneficial to survival and neurological outcomes following cardiac arrest. However, benefit of TTM over non-TTM might be diminished along with improvement in post-cardiac arrest care.

## Declarations

### Ethics approval and consent to participate

This retrospective cohort study was approved by the Institutional Review Board of NTUH (202111079RINA) and waived participant consent.

### Consent for publication

not applicable

### Availability of data and material

The data sets used and analysed during the current study are available from the corresponding author on reasonable request

### Authors’ contributions

All authors contributed to the study conception and design. Material preparation and data collection were performed by CWT and OHN. The analysis was performed by TMS, TYT and WCH. The first draft of the manuscript was written by CWT. TMS, HCH and CWJ revised it critically for important intellectual content.

## Data Availability

All data are available in the manuscript.

## Acknowledgements

The authors would like to express their thanks to the staff of National Taiwan University Hospital-Statistical Consulting Unit (NTUH-SCU) for statistical consultation.

